# The non-coding *GBA1* rs3115534 variant is associated with REM sleep behavior disorder in Nigerians

**DOI:** 10.1101/2023.11.07.23298092

**Authors:** Oluwadamilola O. Ojo, Sara Bandres-Ciga, Mary B. Makarious, Peter Wild Crea, Dena G. Hernandez, Henry Houlden, Mie Rizig, Andrew B. Singleton, Alastair J. Noyce, Mike A. Nalls, Cornelis Blauwendraat, Njideka U. Okubadejo, the Nigeria Parkinson’s Disease Research Network and the Global Parkinson’s Genetics Program (GP2)

## Abstract

**Background:** Damaging coding variants in *GBA1* are a genetic risk factor for rapid eye movement sleep behavior disorder (RBD), which is a known early feature of synucleinopathies. Recently, a population-specific non-coding variant (rs3115534) was found to be associated with PD risk and earlier disease onset in individuals of African ancestry.

**Objectives:** To investigate whether the *GBA1* rs3115534 PD risk variant is associated with RBD.

**Methods:** We studied 709 persons with PD and 776 neurologically healthy controls from Nigeria. The *GBA1* rs3115534 risk variant status was imputed from previous genotyping for all. Symptoms of RBD were assessed with the RBD screening questionnaire (RBDSQ).

**Results:** The non-coding *GBA1* rs3115534 risk variant is associated with possible RBD in individuals of Nigerian origin (Beta = 0.3640, SE = 0.103, P =4.093e-04), as well as after adjusting for PD status (Beta = 0.2542, SE = 0.108, P = 0.019) suggesting that this variant may have the same downstream consequences as *GBA1* coding variants.

**Conclusions:** We show that the non-coding *GBA1* rs3115534 risk variant is associated with increased RBD symptomatology in Nigerians with PD. Further research is required to assess association with polysomnography-defined RBD.

## Introduction

Glucocerebrosidase (*GBA*1) variants have been documented as the most significant genetic risk for Parkinson’s disease (PD) globally.^1^ *GBA1*-associated PD has an earlier age of disease onset, faster motor progression and more frequent non-motor symptoms specifically cognitive decline/dementia, visual hallucinations, hyposmia, autonomic features and rapid eye movement (REM) sleep behavior disorder (RBD). ^2, 3^

One of the early clinical symptoms or prodromal features of synucleinopathies is RBD and over 80% of patients with this sleep disorder will ultimately be diagnosed with PD, dementia with Lewy bodies (DLB), or in rare cases, multiple system atrophy (MSA) within 10–15 year.^4, 5^ Currently, RBD stands as a powerful indicator for the emergence of dementia in individuals with PD, and it is linked to a faster disease progression.^6^ RBD occurs in approximately 50-80% of all DLB cases and 30–60% of PD patients.^7, 8^

The first genome-wide association study (GWAS) in RBD recently nominated two coding variants at the *GBA1* locus, including p.Glu365Lys (rs2230288) and p.Asn409Ser (rs76763715), as linked to RBD risk in the European ancestry population (p.Glu326Lys; OR = 2.09, 95% CI = 1.73–2.54, P= 4.87E-14; p.Asn370Ser; OR = 2.84, 95% CI = 2.06–3.92, P= 1.68E-10).^9^ To the best of our knowledge, no research aimed at exploring genetic risk factors linked to RBD in the African population has been performed.

The first GWAS of PD in the African and African admixed populations has identified a novel population-specific intronic variant linked to PD risk and age at onset at the *GBA1* locus as the major genetic contributor to PD in this population (risk, rs3115534-G; OR=1.58, 95% CI = 1.37 - 1.80, P=2.397E-14; age at onset, BETA =-2.004, SE =0.57, P = 0.0005).^10^ This variant exerts substantial risk of PD as compared with common variation identified through GWAS and it was found to be present in approximately 55% of the PD cases of Nigerian ancestry and 39% of the PD cases from African admixed ancestry.^10^ Importantly, all current *GBA1* knowledge, ranging from clinical to functional, is based on *GBA1* coding variants and not much is known about the impact of non-coding variants on disease etiology. Here, we aimed to investigate the role of *GBA1* rs3115534 and the risk of RBD in the African population.

## Methods

### Study design

This is a secondary analysis of data obtained from study participants recruited by neurologists from the Nigeria Parkinson’s Disease Research (NPDR) network.^11^ Research ethics approval for the study was obtained from the institutional health research ethics committees, the National Health Research Ethics Committee (NHREC) in Nigeria and the University College London (UCL) Institutional Review Board. All participants provided written informed consent. Individuals with PD fulfilled the United Kingdom Parkinson’s Disease Society Brain Bank (UKPDSBB) criteria (Hughes et al. 1992).^12^ Controls were healthy volunteers of Nigerian origin, with no known family history of PD, no clinically evident neurological condition, and matched for age.

### Clinical data collection

The RBD screening questionnaire (RBDSQ), a specific questionnaire for RBD, was used to assess the most prominent clinical features of RBD.^13^ It is a 10-item, patient self-rating instrument with short questions to be answered by either ‘yes’ or ‘no’. The validity of the questionnaire has been shown to perform with high sensitivity and reasonable specificity in the diagnosis of possible RBD (pRBD).^13,14^ Briefly, items 1 to 4 address the frequency and content of dreams and their relationship to nocturnal movements and behavior. Item 5 asks about self-injuries and injuries of the bed partner. Item 6 consists of four sub items assessing nocturnal motor behavior more specifically, e.g., questions about nocturnal vocalization, sudden limb movements, complex movements, or bedding items that fell down. Items 7 and 8 deal with nocturnal awakenings. Item 9 focuses on disturbed sleep in general and item 10 on the presence of any neurological disorder. The maximum total score of the RBDSQ is 13 points.

### Genotyping

DNA extraction as previously described was done either from saliva samples collected using DNA Genotek saliva kits or from venous whole blood samples using standard protocols while genotyping was performed at the National Institutes of Health/Laboratory of Neurogenetics using the NeuroBooster Array (https://github.com/GP2code/Neuro_Booster_Array).^10^

### Imputation procedures

Genetic data were QCed and imputed using standard GP2 pipelines and protocols (https://github.com/GP2code/). As previously reported, the *GBA1* rs3115534 was reliably imputed in all samples.^10^

### Statistical analysis

All samples with complete data were included in the analyses. Two initial screening models were built to test the association between the quantitative RBD score and a dichotomized pRBD outcome with the outcome positive at RBD score six or higher. For the linear regression model testing RBD score as the outcome and the logistic regression model testing pRBD status as the outcome, female sex and enrollment age were used as covariates with the dosage of the *GBA1* rs3115534 variant of interest as the exposure. These models were subsequently repeated with an additional adjustment for PD case status. These same models were then subset to only PD cases and only neurologically normal controls. While the control analyses maintained the original set of covariates, the PD case-only analysis added case age at onset (instead of enrollment age) and disease duration as co-variates.

## Results

In total we included 709 PD cases and 776 neurologically healthy controls of Nigerian descent and assessed whether the novel *GBA1* non-coding rs3115534 risk variant had an effect on pRBD status. Similar to previous observations, the *GBA1* non-coding rs3115534 risk variant was overrepresented in the PD cases vs the neurologically healthy controls (MAF_cases = 0.34 vs MAF_controls = 0.21). A detailed description of demographics and clinical characteristics of the participants under study can be seen in **Table 1**. As expected, PD cases had a higher frequency of pRBD (score >=6) compared with controls (28.2% vs 6.6%, Chi-square value = 121.95, P= 2.37E-28). Initial screening models combining cases and controls showed significant associations between the *GBA1* non-coding rs3115534-G allele and RBD status (Beta = 0.3640, SE = 0.103, P =4.093e-04) as well as the continuous RBD score (Beta = 0.5509, SE = 0.116, P = 2.043e-06). The association between RBD score and GBA1 non-coding rs3115534-G allele persisted after adjusting for PD case status (Beta = 0.2542, SE = 0.108, P = 0.019), showing that the association with *GBA1* rs3115534-G and severity of pRBD is at least partially independent of PD case status in this dataset. The association with RBD score persists when data is subset to only cases (Beta = 0.4166, SE = 0.174, P = 0.017). There are no associations between either RBD score or RBD status among neurologically normal controls (P > 0.05). For a detailed description of all statistical tests performed see **Table 2**.

**Table 1:** Demographics and clinical characteristics of the Nigerian cohort under study.

|  | **Cases (n=709)** |  |  | **Controls (n=776)** |  |  |
| --- | --- | --- | --- | --- | --- | --- |
| Sex | F:195<br>M:514 |  |  | F:265<br>M:511 |  |  |
| Age at onset for cases and at study for controls | 59.7 (range 17-93) |  |  | 63.4 (range 25-98) |  |  |
| RBD average score (SD) | 4.19 (3.25) |  |  | 1.74 (2.05) |  |  |
| RBD status $\geq 6$ (percentage of total) | 200 (28.2%) | | | 51 (6.6%) | | |
| *GBA1* rs3115534 carriers | GG (n=101) | GT (n=285) | TT (n=323) | GG (n=34) | GT (n=298) | TT (n=444) |
| RBD average score (SD) | 4.55 (3.20) | 4.44 (3.31) | 3.85 (3.18) | 1.35 (2.10) | 1.87 (2.16) | 1.70 (1.97) |
| RBD status $\geq 6$ (percentage of total) | 31 (31%) | 90 (32%) | 79 (24%) | 1 (3%) | 24 (8%) | 26 (6%) |

**Footnote:** RBD = Rapid Eye Movement (REM) Sleep Behavior Disorder, F=Female, M=Male, SD = standard deviation

**Table 2:** Regression analyses detailing parameter estimates associated with *GBA1* rs3115534.

| Outcome | Samples | Covariates | Beta | Standard error | P-value (two-sided) |
| --- | --- | --- | --- | --- | --- |
| RBD score | All | Standard | 0.5509 | 0.116 | **2.043E-06** |
| RBD score | All | Standard with additional adjustment for PD status | 0.2542 | 0.108 | **0.019** |
| RBD score | PD cases | Modified to adjust for AAO and disease duration | 0.4166 | 0.174 | **0.017** |
| RBD score | Neurologically normal controls | Standard | 0.0258 | 0.126 | 0.838 |
| RBD status | All | Standard | 0.3640 | 0.103 | **4.093E-04** |
| RBD status | All | Standard with additional adjustment for PD | 0.1912 | 0.106 | 0.071 |
| RBD status | PD cases | Modified to adjust for AAO and disease duration | 0.2034 | 0.118 | 0.084 |
| RBD status | Neurologically normal controls | Standard | 0.1014 | 0.244 | 0.678 |

Footnote: Standard covariates included sex and enrollment age. RBD scores ranged from 0-13, RBD status = dichotomous trait with 0 being RBD score <6 and 1 being RBD score ≥6.

## Discussion

Identifying RBD symptoms in the general population becomes especially important when considering its robust connection to alpha-synucleinopathies including PD, DLB and MSA. Because RBD can manifest many years before the onset of obvious motor symptoms, exploring genetic risk factors for RBD are crucial for prompt detection and intervention or disease modification.

Here we perform the first genetic assessment of RBD risk in the Nigerian population. Our results demonstrate the association between the novel *GBA1* rs3115534 PD risk variant and an increased risk of RBD symptoms, and show that risk carriers are ∼1.43 times more likely to have RBD symptoms than non-carriers. Our results are in line with those from previous PD studies, including other ethnicities, which clearly demonstrated similar relationships between *GBA1* coding variants and the risk for RBD. ^15^ The fact that RBD serves as a potent predictor for the onset of dementia suggests that this variant may also be implicated in PD-dementia and DLB risk. In the current study, we were unable to explore whether this variant is associated with dementia risk in the African population given that we did not have access to dementia data. Additionally, the most commonly used cognitive screening tests such as MoCA and MMSE do not seem to be well suited for this population. ^16^ (Rossetti et al. 2017).

Our study has some limitations. First, questionnaires are subject to recall bias and inaccuracies. Validation of current scales are necessary to ensure reproducibility, reliability and accuracy as polysomnography-defined RBD data becomes available. In the present study, we were not able to confirm that RBD symptoms preceded PD diagnosis due to the lack of longitudinal data in this cohort. Further studies are needed to explore whether *GBA1* rs3115534 affects the rate of phenoconversion from RBD to PD and other synucleinopathies such as DLB and MSA.

In summary, we show here that the non-coding *GBA1* rs3115534 is associated with symptoms of RBD defined by a commonly used screening questionnaire. Identifying individuals at risk of RBD represent an appealing target for clinical trials focused on preventing neurodegeneration in conditions such as PD and DLB. As neuroprotective treatments emerge in the future, an early identification of RBD will be key to treat these conditions.

## Data Availability

All data used in this manuscript is available via GP2 at the https://www.amp-pd.org/portal.

https://www.amp-pd.org/

https://1drv.ms/x/s!Aj-0YTgBXcwfhJ9orGkbfhHqocIZEA

https://docs.google.com/spreadsheets/d/1dum-N2jBjW8DUzRjg2z_VGgJ0WPsSOZ8/edit#gid=1769939610

## Code availability

All code is available as follows: https://github.com/GP2code/GBA1_RBD_NIGERIA; Zenodo: https://zenodo.org/record/8399986

## Data availability

All data used in this manuscript is available via GP2 at the https://www.amp-pd.org/ portal.

## Acknowledgements

This work was supported in part by the Intramural Research Program of the National Institute on Aging (NIA), and the Center for Alzheimer’s and Related Dementias (CARD), within the Intramural Research Program of the NIA and the National Institute of Neurological Disorders and Stroke. Data used in the preparation of this article were obtained from Global Parkinson’s Genetics Program (GP2). GP2 is funded by the Aligning Science Across Parkinson’s (ASAP) initiative and implemented by The Michael J. Fox Foundation for Parkinson’s Research (https://gp2.org). This work utilized the computational resources of the NIH HPC Biowulf cluster (https://hpc.nih.gov).

## Author roles

Conceptualization and design: O.O., S.B-C, M.B.M. P.W., C.B, M.A.N., A.N., N.U.O Data acquisition, analysis or interpretation: All authors. Drafting of manuscript: O.O., S.B-C, M.B.M., P.W.C., C.B., A.N., N.O. Critical revision of manuscript for intellectual content: All authors. Statistical analysis: M.A.N., S.B-C., M.B.M., C.B. Obtained funding: N.O., M.R., H.H., J.H., A.S.

## Supplemental Files

Supplemental Table 1 – Nigeria Parkinson Disease Research (NPDR) network members https://1drv.ms/x/s!Aj-0YTgBXcwfhJ9orGkbfhHqocIZEA

Supplemental Table 2 - Global Parkinson’s Genetics Program (GP2) members https://docs.google.com/spreadsheets/d/1dum-N2jBjW8DUzRjg2z_VGgJ0WPsSOZ8/edit#gid=1769939610

